# Impact of treatment of varicose veins with polidocanol foam compared to conventional surgery on quality of life

**DOI:** 10.1101/2023.08.25.23294616

**Authors:** Lissa Severo Sakugawa, Felipe Soares Oliveira Portela, Andressa Cristina Sposato Louzada, Maria Fernanda Cassino Portugal, Marcelo Passos Teivelis, Cynthia de Almeida Mendes, Alexandre Fioranelli, Nelson Wolosker

## Abstract

**Background and objective:** Lower limb varicose veins are very prevalent and there are several treatment options available, including conventional surgery and polidocanol foam sclerotherapy. Few studies analyze therapeutic modalities based on PROMs (patient-reported outcome measures). The aim of this study was to evaluate the impact of treatment with polidocanol foam sclerotherapy compared to conventional surgery in a large sample, based on an analysis of PROMs.

**Methods:** This was a prospective, observational, and qualitative study of 205 patients who underwent varicose vein treatment with polidocanol foam sclerotherapy (57 patients - 90 legs) or conventional surgery (148 patients - 236 legs). Patients were assessed preoperatively and 30 days after the procedure using venous disease severity scores (VCSS) and specific venous disease quality of life questionnaires (VEINES-QoL/Sym).

**Results:** Both treatments promoted a significant improvement in VCSS and VEINES 30 days after the procedure (p < 0.05). However, surgery promoted a greater improvement in VCSS (on average 4.02 points difference, p < 0.001), VEINES-QoL (8 points difference, p < 0.001) and VEINES-Sym (on average 11.66 points better, p < 0.001) compared to sclerotherapy. Post-operative pain and aesthetic concern about the legs were the domains of the questionnaires that had the greatest impact on this difference between the two types of treatment, generating worse results with sclerotherapy.

**Conclusion:** Polidocanol foam sclerotherapy and conventional surgery have a positive impact on quality of life after 30 days, but there is a more significant improvement in patients undergoing conventional surgery.

## INTRODUCTION

Lower limb varicose veins (VV) are permanently dilated and tortuous veins that affect up to 50% of men and 70% of women. They can cause from mild symptoms such as fatigue, feeling of heaviness or burning, swelling, and itching in the legs, to more serious complications such as phlebitis, ulcers, and varicose veins bleeding^1^.

Due to its high prevalence, the number of surgeries for the treatment of VV is also high, reaching 80,000 per year in England and 70,000 per year in Brazil^2,3^. The conventional surgical approach is the most widely used treatment in clinical practice with excellent results, which is why this is the preferred method in the Brazilian public health system. Other approaches such as the injection of sclerosing substances and laser or radiofrequency ablative techniques are also alternatives^4^.

Assessing the improvement of VV symptoms is complex, as they are not pathognomonic, are poorly correlated with anatomical and imaging parameters and strongly associated with psychological aspects^5^. A strong correlation between quality of life and the clinical results of surgical treatment for VV has already been demonstrated^6^. In recent decades, for various health conditions (and also for varicose veins), healthcare systems have increasingly recognized the importance of using patient-centered tools to offer better treatments in terms of quality, efficacy, and cost-effectiveness^7^. PROMs (patient-reported outcome measures) are instruments that allow the collection of information on symptoms, functional status, and quality of life, and make it possible to define the success of therapeutic interventions based on patient experience^8^.

However, in the scientific literature, the definition of success in treating VV is still fundamentally based on technical criteria^9,10^. Few studies are based on the analysis of PROMs to define the effectiveness of treatments, and several of them use generic questionnaires that are not specific to venous disease^11,12^. Also, few studies assess quality of life with the use of polidocanol^13,14^. In addition, most studies include few (if any) patients with more advanced degrees of chronic venous disease (CVD)^14,15^. There are no impact studies carried out in low- and middle-income countries using PROMs.

This study aims to evaluate the impact of treating CVD with polidocanol foam sclerotherapy (PFS) compared to conventional surgery (CS) in a large sample (205 patients) using PROMs.

## METHODS

### Study design

Prospective, observational, and qualitative study with 205 patients undergoing treatment for VV, 57 of whom underwent polidocanol sclerotherapy (foam group), and 148 underwent conventional surgical treatment (surgical group), considered the control group, in 2 institutions of the public health system in the city of São Paulo - Brazil, from October/2021 to October/2022. Patients were included consecutively.

The research was approved by the institutions’ ethics committees (protocols: CAAE 51668821.6.0000.0071 and 51668821.6.3001.0083). Consent Form was applied to all patients who agreed to take part in the study.

Patients undergoing PFS were treated on an outpatient basis. The tributary veins were punctured under direct vision, with the application of 1% polidocanol foam. In the case of insufficient saphenous veins, ultrasound-guided distal puncture of the saphenous vein was performed, with injection of 3% polidocanol foam.^16^.

Patients undergoing CS were treated in an operating room under spinal anesthesia. VV were resected through staggered incisions. In cases of great saphenous vein insufficiency, treatment was performed through an incision at the root of the thigh, with ligation of tributary veins and resection of the insufficient segment with a pin-stripper.

Each individual was assessed on two occasions: preoperatively and 30 days after treatment. On each of these occasions, the CEAP classification (Clinical manifestations, Etiologic factors, Anatomic distribution of disease, Pathophysiologic findings) was assessed and the Venous Clinical Severity Score (VCSS)^17^ and VEINES/QOL-Sym^18^ (Portuguese version)^19^ questionnaires were applied.

Regarding the CEAP, we basically assessed clinical manifestations (C), which stratifies the manifestations of venous disease into 7 categories:

- C0: No signs of venous disease
- C1: Telangiectasies or reticular veins
- C2: Visible varicose veins
- C3: Edema
- C4: Skin changes - hyper-pigmentation, eczema, lipodermatosclerosis
- C5: Healed ulcer
- C6: Active ulcer

VCSS comprises 10 attributes (pain, varicose veins, edema, pigmentation, inflammation, induration, number of ulcers, duration of ulcers, size of ulcers, compressive therapy) which are rated from 0 to 3 (absent, mild, moderate, severe)^20^. All these items were analyzed in this study.

VEINES/QOL-Sym is a quality-of-life assessment tool that is specific for venous disease and consists of 26 items, including questions about symptoms (10 items), limitation of daily activities (9 questions), psychosocial impact (5 items), change in complaints over a period of 1 year (1 item) and the time of day when symptoms are most frequent^21^.

In order to better analyze the variations in quality of life before and after treatment, the VEINES scores were converted into a 0-100 scale, using a method previously described in the literature^22^. All items of the originally described questionnaire were analyzed preoperatively and 30 days after each type of procedure.

Demographic data of patients undergoing CS or PFS was analyzed, as well as the preoperative CEAP classification. VCSS and VEINES/QOL-Sym scores were calculated preoperatively and postoperatively, with comparative analyses between the periods and between the two treatment groups.

The factors (within each score) that most influenced the differences between the treatments were also analyzed. For this purpose, the average variation of each score was calculated for each type of procedure, with linear regression analysis to define the items of each questionnaire that proved to be statistically significant.

### Statistical analysis

It was carried out using SPSS software version 22 (IBM - Armonk, New York, USA). The Student’s t-test was used to assess continuous variables, while the chi-square test was used to analyze categorical variables.

In order to analyze the variations in quality of life scores at each time point (pre- and post-treatment), generalized estimating equation with a normal distribution and identity link function was used, assuming an AR(1) correlation matrix between the time points and Bonferroni multiple comparisons. Multiple linear regression was used in the multivariate analysis.

For all tests, a P-value less than or equal to 0.05 was considered statistically significant.

## RESULTS

In all, 57 patients (90 limbs) were treated with PFS, and 148 patients (total of 236 limbs) underwent CS.

Demographic data for the two groups are presented in Table 1. We observed that patients undergoing PFS were significantly more hypertensive (58.8% vs 29.1%, p < 0.001), diabetic (29.4% vs 8.1%, p < 0.001) and obese (48% vs 24.2%, p < 0.001). Females were predominant in both groups, with no significant difference (p = 0.121) between them. Patients in the foam group were significantly older (p = 0.002) and had a higher body mass index (p = 0.002).

**Table 1:**
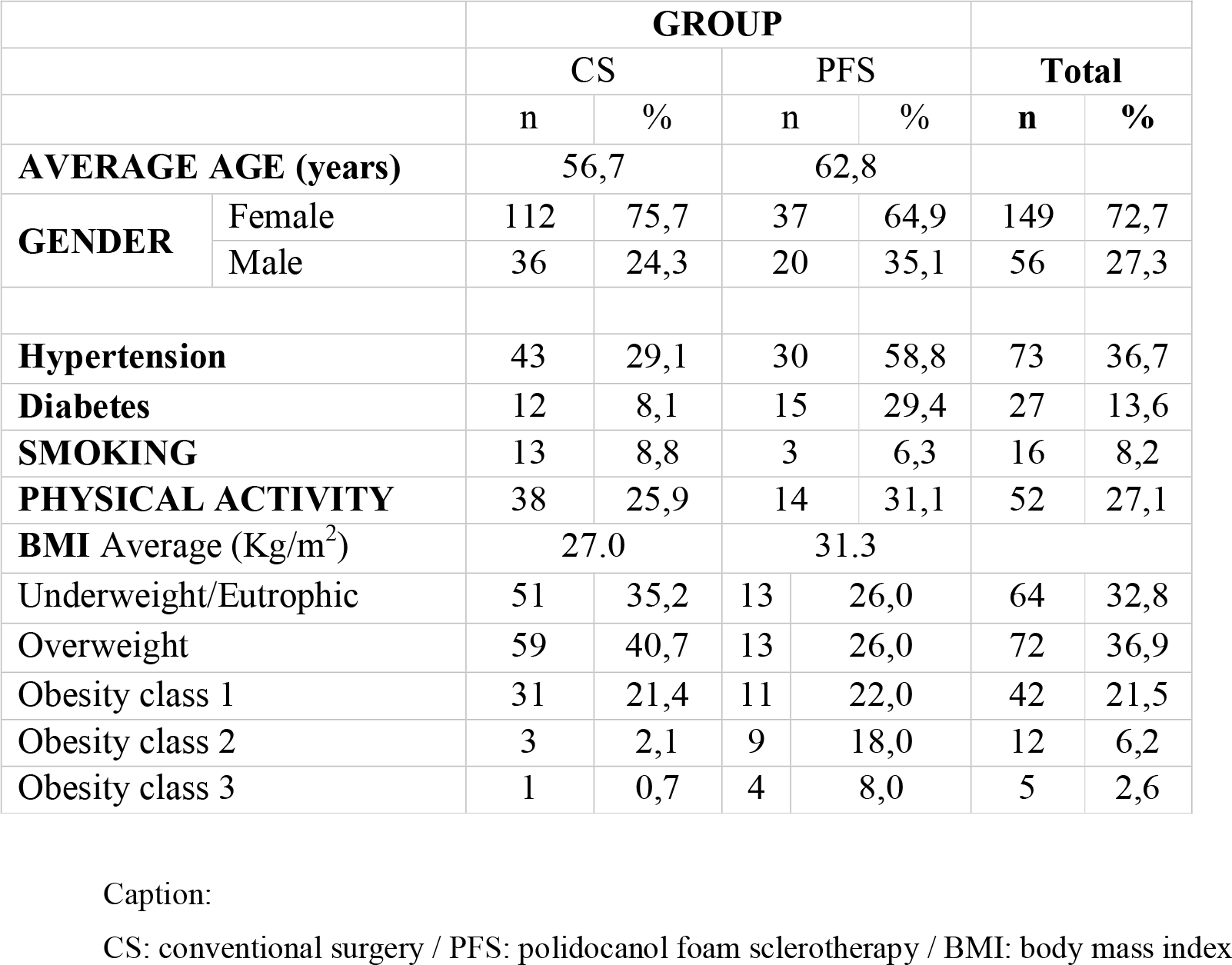
demographics and clinical characteristics.

The CEAP classification (of the treated limbs) is detailed in Table 2. Although the foam group had a higher proportion of patients with more advanced degrees of CVD (24.4% of individuals with CEAPs C5 and C6, compared to 9.3% of patients in the control group), there was no statistically significant difference between the groups (p = 0.304).

**Table 2:** preoperative CEAP classification of treated limbs.

|  | PFS | CS |
| --- | --- | --- |
| C1 | 7 (7,7%) | 0 |
| C2 | 18 (20,0%) | 38 (16,1%) |
| C3 | 23 (25,5%) | 60 (25,4%) |
| C4 | 20 (22,2%) | 116 (49,1%) |
| C5 | 9 (10,0%) | 0 |
| C6 | 13 (14,4%) | 22 (9,3%) |
| Total | 90 | 236 |

Caption: CS: conventional surgery / PFS: polidocanol foam sclerotherapy

A comparison of the scores before and after treatment, for conventional surgery and sclerotherapy, is shown in Table 3:

**Table 3:** Pre- and post-treatment scores (mean ± standard deviation):

|  | CS | PFS |
| --- | --- | --- |
| VCSS pre | 18,04 ± 4,52 | 20,33 ± 6,53 |
| VCSS post | 14,48 ± 2,66 | 18,50 ± 7,09 |
| VEINES-QoL pre | 64,62 ± 11,12 | 64,48 ± 11,02 |
| VEINES-QoL post | 80,09 ± 9,29 | 71,99 ± 11,28 |
| VEINES-Sym pre | 61,93 ± 15,78 | 59,29 ± 15,57 |
| VEINES-Sym post | 82,11 ± 10,56 | 70,50 ± 14,80 |

Caption: CS: conventional surgery / PFS: polidocanol foam sclerotherapy / pre: scores pre-treatment / post: scores post-treatment

Before treatment, patients undergoing PFS had a significantly worse mean VCSS than those undergoing CS (20.33 vs 18.04, p = 0.019). There was no difference between the groups in the pre-procedure assessment in relation to the VEINES scores for quality of life (control 64.62 vs foam 64.48, p > 0.999), nor for symptoms (control 61.93 vs foam 59.29, p > 0.999).

Both treatments led to a significant improvement in scores 30 days after the procedure (p ≤ 0.05 in all scenarios), but after the treatments, the surgical group showed a higher rate of improvement than the foam group. The post-treatment VCSS of patients undergoing CS was on average 4.02 points lower (better) than that of patients undergoing PFS (14.48 vs 18.50, p < 0.001).

Regarding VEINES, patients in the surgical group scored an average of 8 points better than those in the foam group in terms of quality of life (80.09 vs 71.99, p < 0.001) and an average of 11.66 points better in terms of symptoms (82.11 vs 70.50, p < 0.001).

Next, we analyzed all criteria in each questionnaire to determine which factors most influenced the better performance of the surgery compared to the foam treatment. Pain was the aspect that most contributed to significantly better post-operative VCSS in the control group compared to the foam group. Although the groups were initially similar to each other regarding this symptom (p > 0.999), patients who underwent CS had a significant improvement in this symptom 30 days after the procedure (mean improvement in VCSS of 1.28 points, p < 0.001), while individuals who underwent PFS did not have a statistically significant improvement (mean improvement in VCSS of 0.5 points, p = 0.642).

In VEINES-Sym, the domains of edema and sensation of heat or burning were those that did not improve significantly in the foam group and contributed to worse scores in postoperative evaluation compared to the control group. Regarding edema, there was a mean difference of 0.95 points (p < 0.001) in the postoperative score in favor of the surgical group. As for the burning sensation, there was an average improvement of 1.31 point in surgical group (p < 0.001), compared to a small, non-significant improvement (0.24 points, p > 0.999) in the foam group.

Regarding VEINES-QoL, concern about the risk of falling and the influence of leg appearance on clothing choice were the factors that made the post-procedure evaluation worse in the foam group compared to the control group. CS led to an average improvement of 1.78 points in individuals’ concern about the risk of falling (p < 0.001), but there was a slight non-significant worsening of 0.16 points (p > 0.999) after PFS. As for the influence of leg appearance on clothing choice, surgery promoted a significant average improvement (2.05 points, p < 0.001) while foam promoted a slight non-significant improvement (0.45 points) (p > 0.999).

In multivariate analysis, for the VCSS, a higher preoperative (baseline) score contributed to a worse post-treatment result for the foam group compared to the control group (p < 0.001). In the VEINES, grade 2 obesity influenced the foam group’s worse results 30 days after the procedure, both in terms of symptoms and quality of life (p < 0.001 in both scenarios).

## DISCUSSION

Although interventions to treat CVD are among the most commonly performed in clinical practice, the evaluation of their results is still inconsistent^23^. Most of the studies available in the literature evaluate the success of VV treatment based on technical criteria (presence of venous reflux, remaining veins), with little emphasis on clinical criteria related to the symptoms and quality of life of individuals.^24,25^.

Quality of life questionnaires and PROMs provide a tool for assessing the impact of VV or their treatment on patients’ quality of life^26^. There is no perfect score that allows us to assess all the clinical complexity and patient perception of venous disease. In this study, for a more complete analysis, we decided to use a score based on clinical evaluation (VCSS) associated with one that assesses patients’ perceptions of their symptoms (VEINES-Sym) and the extent to which they interfere with their quality of life (VEINES-QoL).

VCSS makes it possible to measure subtle changes in the severity of CVD, but it encompasses many symptoms that are not pathognomonic of VV and may not be very specific, especially for lower CEAPs (C1 to C3). VEINES, on the other hand, is currently one of the most acceptable options when it comes to assessing the impact on quality of life of the whole spectrum of CVD^27^. However, in the original VEINES calculation, there may be a significant limitation in evaluating changes in quality of life between two different moments, which is why we opted to use an alternative form, which adapts this calculation in an objective way, as previously described in the literature^28^.

In this study, 44 legs (13.5%) had active ulcers at the time of treatment. In the international literature, most papers do not include CEAP C6 patients^11,14,15^, which makes this data unique to this study. In Brazil, around 75% of the population (around 160 million individuals) depends exclusively on the care provided by the public health system. Patient access to health services often takes a long time, especially for non-urgent diseases such as VV. This delay in getting medical attention may explain the higher proportion of individuals with more advanced degrees of venous disease^29^.

Although varicose vein surgery represents a low risk for patients, sclerotherapy is an even less invasive alternative, as it does not require hospitalization or invasive procedures^30^. In our series, we found that patients chosen for treatment with PFS were older than those who underwent CS, which is consistent with other available studies^11,15^. In general, studies also show an improvement in quality of life immediately after interventions, both with sclerotherapy and conventional surgery^11,13,14^. However, what is observed in large trials with long-term follow-up is that there is a late deterioration (usually after 2 years of treatment) in clinical severity and quality of life scores^12,15^ as CVD recurs. In our sample, we observed an improvement in quality of life in both patients undergoing PFS and those undergoing CS, although post-operative improvement was more significant in patients undergoing surgery. This result can be explained by effects of the procedures themselves and by characteristics of the patients chosen for either treatment.

In our study, pain was the factor that had the greatest impact on a worse post-treatment outcome in the foam group. This finding is not compatible with other studies, which point to more pain after treatment with conventional surgery^12,31^. Perhaps the greater perception of pain after sclerotherapy in our sample could be related to the effects of polidocanol, which can cause phlebitis due to the local inflammatory reaction^32^.

In addition, phlebitis and hyperpigmentation possibly associated with foam application^13^ may correlate with the worse results observed in the VEINES quality of life domain after the procedure, influencing patients’ concern about the appearance of their legs.

Finally, aspects related to the patients’ own characteristics end up having an influence on their personal perception of improvement in the post-operative period. The individuals in the foam group were older and had more comorbidities. These two aspects, in other studies, have already been independently associated with worse quality of life scores specific to varicose veins, regardless of the degree of venous disease^24,33–35^.

It is important to note that, despite these differences, both treatments are effective in dealing with VV. Surgery is associated with positive (early and late) results in technical^12,31^ and clinical aspects^36^. And sclerotherapy, despite the possible need for reoperations and potential local side effects, is also very applicable and cost-effective, as it is a procedure with lower costs, without the need for hospitalization or peri-procedural anesthesia, with an earlier return to activities^37^.

In a context such as the Brazilian public health system, where many patients have multiple comorbidities or are elderly, factors which may make it difficult or impossible to perform surgery, sclerotherapy is a viable and effective alternative.

The limitations of this study were that, although consecutive patients were included, there was no randomization process for one or other type of treatment. As it evaluated patients treated in the public health system, it was only possible to evaluate the treatment options available there, and it was not possible to evaluate other methods currently available, such as ablative techniques.

## CONCLUSION

PFS and CS have an immediate (30-day) positive impact on the quality of life of individuals with VV.

However, the improvement in quality of life caused by surgery is greater than that provided by sclerotherapy, especially due to a greater impression of pain and aesthetic concerns in the case of individuals treated with foam.

## Data Availability

All data produced in the present study are available upon reasonable request to the authors

## Acknowledgements

none.

